# An accuracy study of the YouDiagnose disease predictive model based on a double-validated real-world dataset from the UK

**DOI:** 10.1101/2023.04.17.23288592

**Authors:** Aswini Misro, Vikash Sharma, Naim Kadoglou

## Abstract

Through the utilisation of algorithms and data-driven analytics, predictive technology can be leveraged to provide clinicians with invaluable insight into their patients’ conditions, allowing for more accurate, informed, and timely decision-making in the fast-paced clinical environment. This study aims to provide a preliminary proof of concept with a double-validated real-world dataset from the UK. The YouDiagnose predictive model, developed using retrospective data from over 41,257 patients’ data, was assessed by testing it with a double-validated real-world dataset from the UK and the machine predictions have been compared here with the final diagnosis of the diseases as the gold standard. Out of the total of 433 cases, 60 cases had a mismatch in their prediction, all of them being cancer overdiagnosis, resulting in a lower specificity rate of 84.3%. The combined prediction accuracy at the first prediction level was 86% (n=373) while prediction 1-3 combined was successful in predicting diseases in 93% of the cases when evaluated against the gold standard. The model accurately predicted all 52 cases of cancer, indicating a 100% sensitivity rate. The study shows that the tool can be used in the frontline to accurately screen patients with a high level of confidence in the inclusion of cancer patients. This tool’s high sensitivity means that there is little chance of missing any cancer cases.

## Introduction & scientific justification

The use of data-driven risk analytics in the community has tremendous potential to improve the prospect of early prediction of critical diseases e.g., cancer. By providing risk assessment, disease prediction, automation of triaging, workflow automation and management of care pathways, data-driven risk analytics can enable flagging and early diagnosis of life-threatening diseases such as cancer. This could result in improved patient outcomes and better use of healthcare resources. This data-driven risk analytics is an invaluable tool in the fight against serious diseases.

The benefit to patients and the entire ecosystem is clear. Outpatient waiting times can be significantly reduced leading to more accessible healthcare. Additionally, the quality of interactions between patients and their doctors will be greatly improved, with up-to-date and accurate information providing the basis for a more patient-centric user experience. Clinicians too will experience a reduction in professional burn-out due to the lightening of repetitive and administrative tasks. Please see the discussion section to learn about the use of predictive technology in healthcare.

This pilot study seeks to compare the accuracy of a data-driven model prediction with the final diagnosis backed by investigations such as radiology, pathology and histology. Specifically, this study will analyse the prediction of diseases and the sensitivity of cancer risk prediction. The results of this initial comparison will be invaluable in providing insight into the value of such predictive technologies for front-line triage.

The NHS is facing immense pressure with resource constraints and the backlog of referred cases from the community. Predictive technologies-driven automation can help to improve the system efficiency and cost saving without a huge expense. This technology could enable the NHS to better anticipate demand and manage resources, ensuring that more services can be provided at a much lower cost. With this technological solution, the NHS can rise up to meet the challenges it faces today.

## Ethics approval and consent to participate

At the outset, the research methodology was approved by YouDiagnose Ethical Approval Committee. This research work was commenced after this approval and this study did not involve any non-anonymised human data, tissues, samples, or materials. Consent to participate was obtained from each participant at the beginning of the study. Using the NHS Health Research Authority and United Kingdom Medical Research Council decision-making tool, it was determined that this study would produce generalisable or transferable findings. Therefore, informed consent was obtained from each participant before the structured interview. It was also determined that the study would anonymize the participants to mitigate any risk. All the methodologies were in line with the core practices of the Committee on Publication Ethics (COPE) and the Declaration of Helsinki.

**Figure 1.**
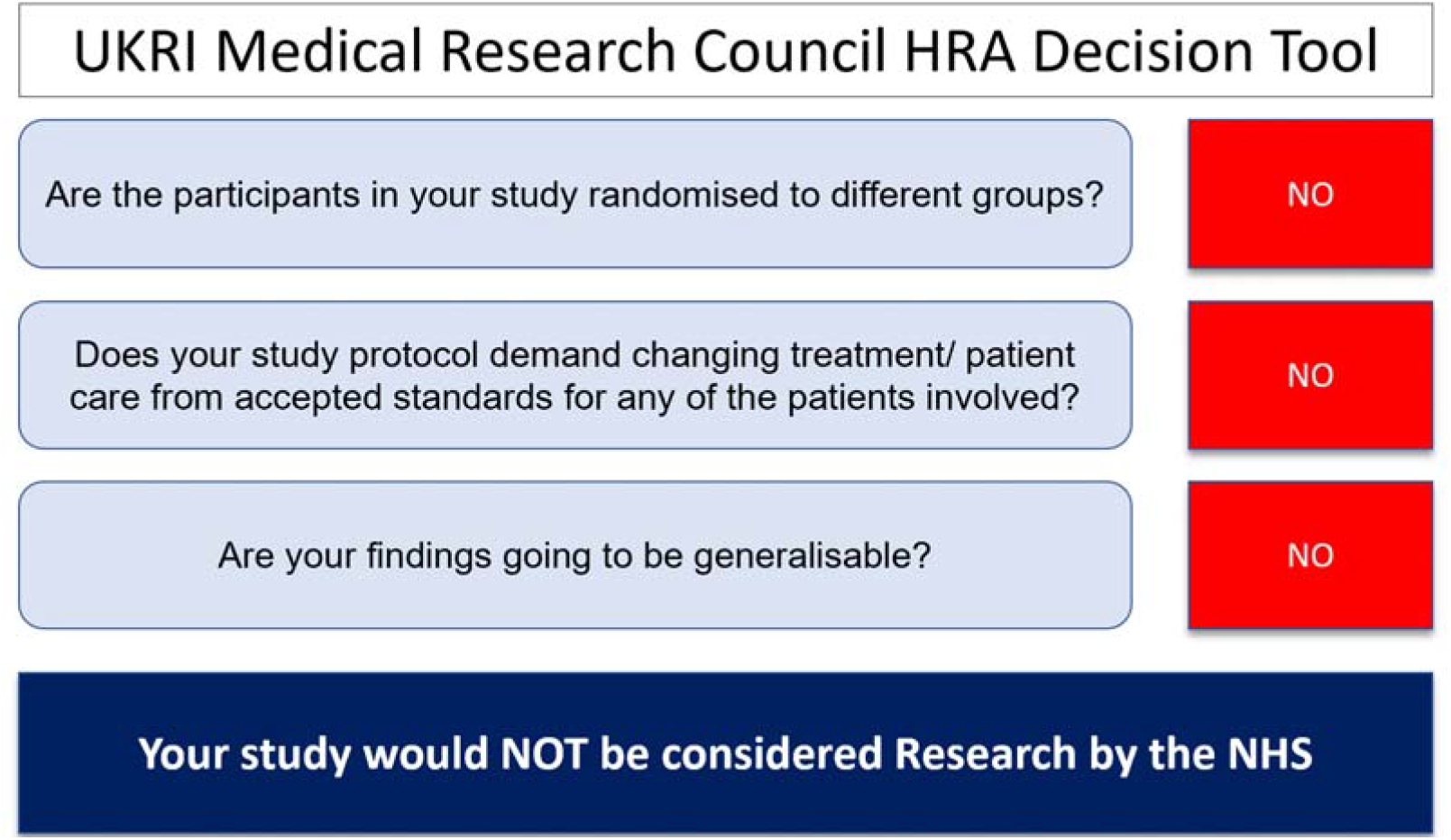
The output of HRA Decision Tool

## Availability of data and materials

It is our policy to protect the intellectual property of our research and development. Therefore, we cannot share the data which was used to build and test the data model. We understand that this may be a disappointment, but we have shared here an example of YouDiagnose’s Federated Autonomous Multi-Disease Deep Learning Database that was used in 2019 and is now defunct. All rights are reserved by YouDiagnose Limited, and the data must not be used for commercial purposes or distribution. The number associated with the data is the case identifier. This is for reference only and the details cannot be disclosed.

## Model design

This model was developed using retrospective data from over 41,257 patients from the Centres of Excellence that practice evidence-based medicine and multidisciplinary disease management. Data were sanitised by data scientists and clinical computer scientists, and 9,109 cases were discarded due to data ambiguity, incomplete data, and data errors. The data underwent double validation, resulting in a high-quality database containing 32037 cases of multi-disease data. The final set was divided into two categories 1. 70% training data and 30% test data.

**Figure 2.**
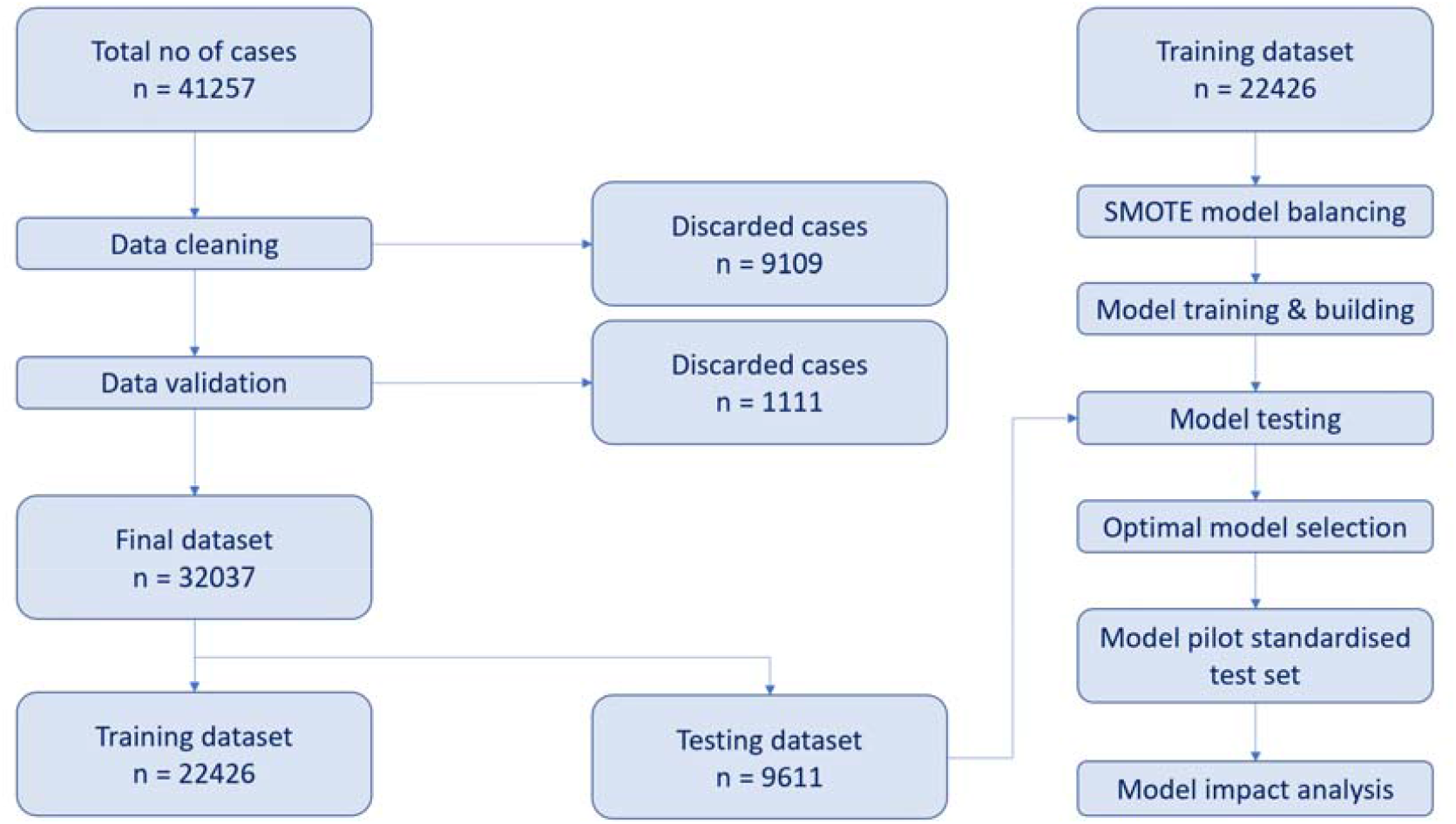
Study & data flow

## Algorithm development

The exploratory data mining approach used unsupervised machine learning in order to develop a relational database and a variety of association rules that interconnect them. As part of this process, the Apriori frequent pattern algorithm was developed to connect various clinical codes used in common day-to-day practice and insurance. This algorithm has proven to be an invaluable tool, offering insight into complex data sets. Other methods were also deployed e.g., XGBoost, and Markov Decision Process for reinforced machine learning to make accurate clinical codes prediction. The latter method helped to use common clinical experiences to integrate into the algorithms e.g., polycystic ovarian disease is commonly associated with infertility. However, it made the dependency on the physician too intense and needed much manual effort to bring accuracy to the model.

### Model building

To remove imbalance effects, a synthetic minority oversampling (SMOTE) method was applied to the training data set. A five-way cross-validated grid search of the training dataset was performed to obtain the best hyperparameters for modelling. Finally, a test data set was used to finally assess the quality of the model. 8 model quality metrics were applied to assess model quality: first prediction accuracy, second prediction accuracy, third prediction accuracy, positive predictive value, sensitivity, specificity, negative predictive values, and AUC (area under ROC).

### Model maturation

For each result, we built models using different ML algorithms, including LR, RF, SVM, K Nearest Neighbor (KNN), lightGBM, XGBoost, and Multilayer Perception (MLP). ML models were run in Python based on the Sklearn library and associated ML modules. The main purpose of the model-building exercise was to predict the most probable disease based on the data provided. One of the priorities was not to miss cancer. Using the exercises above, we built 19 predictive models, each providing a different type of specificity, sensitivity, and positive and negative predictive value. This leads to accurate diagnosis, misdiagnosis, and overdiagnosis.

### Model selection

Results were evaluated by clinical experts in terms of clinical impact. We carried out a real-world and economic impact analysis of these 3 models to study their cost-effectiveness and risk-benefit analysis. The impact analysis of all the 3 models showed the following. The model-1 displayed a relatively high incidence of missed diagnosis, delayed diagnosis, and near-misses. In model - 2, the risk-matrix performance matches with the physician risk assessment and triaging decisions in more than 80% of cases. Model - 3 had a high degree of accuracy; however, it is not cost-effective; on the contrary, it might have many harmful side effects where the potential harm of unnecessary over-diagnosis outweighs any potential benefits.

**Figure 3.**
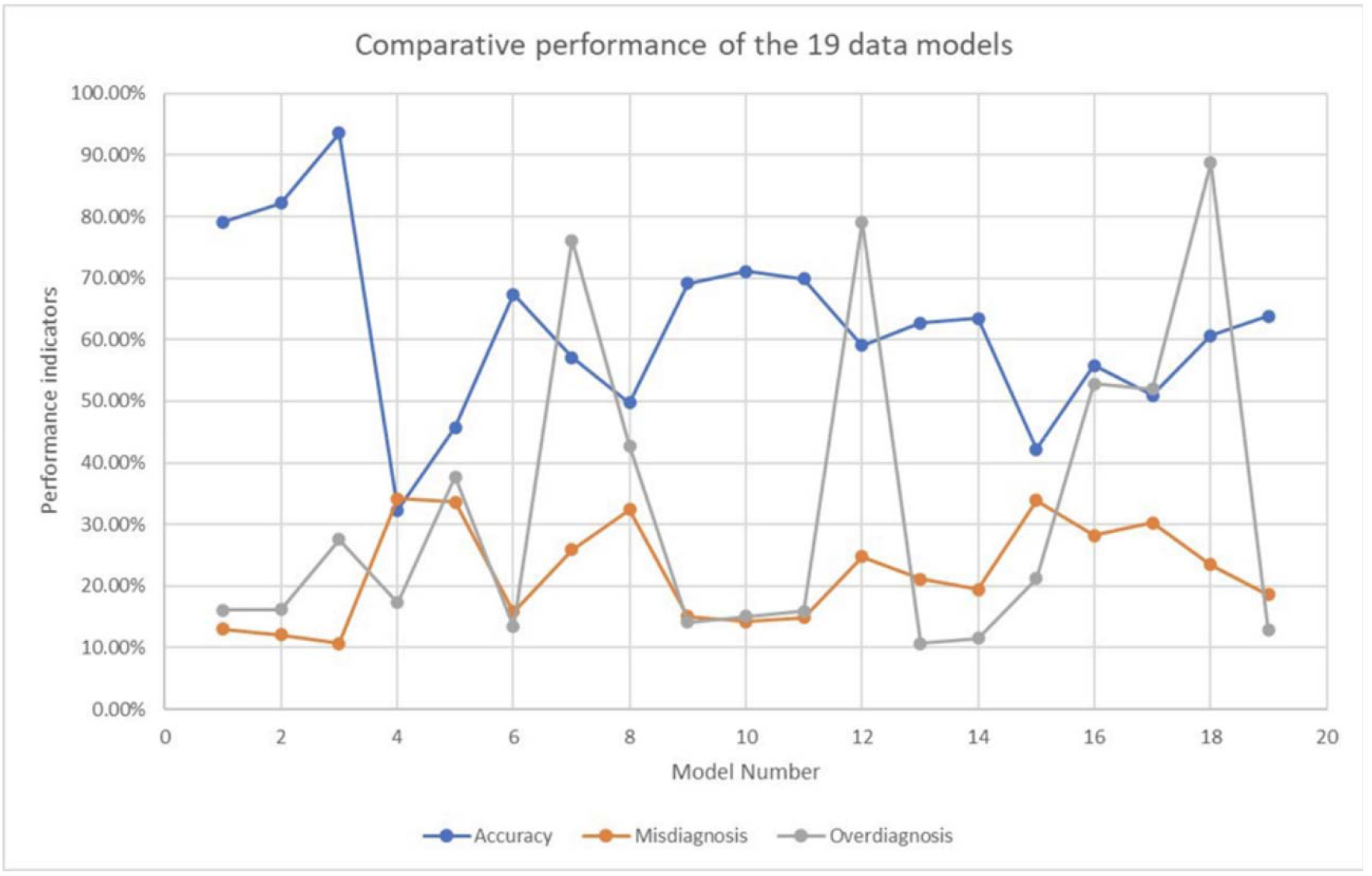
Comparative performance of 19 data models

### Decision Tree Building

Based on the model data points a multi-variate decision tree was designed around various data points to ask the user e.g., patient-relevant questions to generate the data points necessary to build the frontend decision tree of the application

### Real-world dataset preparation

The final model was tested on a double-validated YouDiagnose disease predictive model based on a validated real-world dataset from the UK. Double validation has been performed by specialist surgeons and physicians in relevant fields who have been practising in this field for at least 10 years. They ensured that the case complied with local standards of practice in the UK. However, at times, there are so many ways to investigate and treat the same condition in the same patient that it is impossible to rule out all deviations. The cases with disagreements regarding patient diagnosis and treatment were excluded from the database.

### Evaluation of the test with the real-world dataset

A mismatch (=0) means the model’s prediction didn’t match the gold standard (please see the glossary of terms at the end of the article). Please see Table-1. A perfect match (=2) was considered if the model’s predictions matched the gold standard. If the model prediction matched the gold standard similar disease, it was a similarity match (=1) (please see the glossary of terms at the end of the article).

**Table 1.** showing test results with the real-world dataset.

| Sl No | Parameter | Value | Per cent |
| --- | --- | --- | --- |
| 1 | Total number of cases | 433 | 100% |
| 2 | Total number of mismatch | 60 | 14.0% |
| 3 | Total number of perfect 1st match (A) | 272 | 62.8% |
| 4 | Total number of similarity 1st match (B) | 101 | 23.3% |
| 5 | Total number of 1st prediction match (A+B) | 373 | 86.0% |
| 6 | Total number of overdiagnosis/false +ve cancer | 60 | 14.0% |
| 7 | Total number of missed cancer | 0 | 0.0% |
| 8 | Total number of misdiagnoses | 0 | 0.0% |
| 9 | Total number of 1st prediction match | 272 | 62.8% |
| 10 | Total number of 1st + 2nd prediction match | 363 | 83.7% |
| 11 | Total number of perfect match in 1st + 2nd + 3rd predictions | 403 | 93.0% |

### Findings of the test results

Out of the total of 433 cases, 60 cases had a mismatch in their prediction, with all of them being cancer overdiagnosis. When discussing accuracy at the first prediction level, the number of perfect 1st matches was 62.8% (n=272), while similarity 1st matches stood at 23.3% (n=101). The combined prediction accuracy at the first prediction level was 86% (n=373).

Analysis showed that prediction-1 had a perfect match of 62.8%, meaning that the model was able to predict disease on par with the gold standard in more than 62% of cases. The results, presented in Table-1, show that prediction 1-3 combined was successful in predicting diseases in 93% of the cases when evaluated against the gold standard.

The results of Table-2 demonstrate that the model accurately predicted all 52 cases of cancer, indicating a 100% sensitivity rate. However, the model also generated 60 false positive predictions regarding benign cases, resulting in a lower specificity rate of 84.3%. The overall Positive Predictive Value and Negative Predictive Value were thus impacted accordingly. The Positive Predictive Value (PPV) was 46.4% while the Negative Predictive Value(NPV) was 100%.

**Table 2.** showing model accuracy in cancer prediction.

| Test/Cancer | cancer + | cancer - |
| --- | --- | --- |
| prediction + | 52 | 60 |
| prediction - | 0 | 321 |

| Parameter | Percentage |
| --- | --- |
| sensitivity for predicting cancer | 100.0% |
| specificity for predicting cancer | 84.3% |
| positive predictive value | 46.4% |
| negative predictive value | 100.0% |

### Interpretation

The study shows that the tool can be used in the frontline to accurately screen patients with a high level of confidence in the inclusion of cancer patients. This tool’s high sensitivity means that there is little chance of missing any cancer cases. Furthermore, it can be used to route patients into various levels of care facilities with confidence.

### Limitation

Despite obtaining encouraging results, the current study is limited by the availability of the dataset and its retrospective nature. This may limit its practical value in real-life settings, which warrants further exploration. Testing the tool in a real-life setting in collaboration with senior physicians and surgeons will be extremely beneficial in obtaining an accurate assessment of its real-world performance. This real-world feedback will help refine the model, enabling us to gauge its impact more effectively.

## Discussion

Predictive modelling is a powerful tool in healthcare for predicting medical conditions by analysing patient data. By analysing large amounts of patient data, healthcare providers can create statistical models that can predict the likelihood of a patient having a specific condition. Predictive modelling can also be used to personalize treatment and identify patients at high risk of developing specific medical conditions.

Some of the key benefits of predictive modelling in healthcare include:

- Improved medical decision-making.
- Triaging
- Personalized service provision
- Enhanced operations
- Lower overall costs

Some of the use cases of predictive modelling in healthcare include:

- Forecasting future events to improve the health of individuals.
- Analysing massive amounts of patient, consumer, and market data to output actionable insights about a patient’s future healthcare needs [1]
- Collecting and integrating lifestyle, symptom and treatment data to produce holistic treatment plans [2]

By using these models, frontline workers can better prioritise resources and identify patients who are most at risk of deteriorating while in the hospital. Predictive technologies can help in clinical decision-making, with the objective of improving patient care and reducing healthcare costs. The ultimate goal is to improve patient outcomes while ensuring that the healthcare system is operating as efficiently as possible [3,4,5].

While models can bring speed to medical decision-making, they can never replace the role of human beings in healthcare. Expert professional decisions are vital to personalized care and empathy, something a model can never replicate. However, these models can supplement frontline workers by taking on tasks that require less intuition and more data processing.

For instance, one machine learning application can easily perform the work of tens of human beings. Despite the benefits of technology, it is essential to keep human contact and expert professional decision-making at the forefront of healthcare for optimal outcomes. Predictive technologies are used in various fields such as finance, weather forecasting, and healthcare. In healthcare, predictive analytics solutions rely on big data and artificial intelligence. Predictive analytics in healthcare aggregates vast amounts of patient data incoming from electronic health records (EHR), insurance claims, administrative paperwork, medical imaging, etc. and processes it by searching for patterns. Some examples of predictive analytics in healthcare include detecting early signs of patient deterioration in the ICU and the general ward, suicide, and self-harm prevention, and predicting appointment no-shows [5-11].

## Data Availability

https://predictive-pilot.eniston.com/about-the-study/can-i-see-the-database-which-was-used-to-build-the-model

## Glossary of terms

Similarity match: Similarity match’ term is used when there was a match with a disease which is similar in etiopathogenesis, sharing the presenting features and at least the initial line of treatment. For example, certain types of diseases cannot be distinguished from one another based on presentation, and it is almost impossible to distinguish between them based on medical history alone. Needs scrutiny. However, there are many diseases that share similar aetiology, pathogenesis, symptoms, initial plan, and treatment. Examples can be given of lactation mastitis and breast abscess which are infections involving breast tissue in a breastfeeding woman and need prompt treatment with antibiotics. Both have the same symptoms and cannot be distinguished without ultrasound.
Gold standard: For all cases, the final diagnosis or final diagnosis was accepted as the disease the patient had, as confirmed from the patient’s case record after the completion of investigations and interventions. This was taken as the gold standard for comparing model performance.

## Reference list

1. Alkhaldi, N. (2021). Predictive Analytics In Healthcare: 7 Examples and Risks. [online] ITRex. Available at: https://itrexgroup.com/blog/predictive-analytics-in-healthcare-top-use-cases/.

2. complex, D.B.D.B. is an award-winning writer capable of bridging the gap between, Technology, C.A., innovation and Condition, T.H. (n.d.). How Predictive Modeling in Healthcare Boosts Patient Care. [online] Technology Solutions That Drive Healthcare. Available at: https://healthtechmagazine.net/article/2021/04/how-predictive-modeling-healthcare-boosts-patient-care-perfcon.

3. IBM (2022). What Is Predictive Analytics? | IBM. [online] http://www.ibm.com. Available at: https://www.ibm.com/topics/predictive-analytics.

4. live-hg-ih.cphostaccess.com. (n.d.). Predictive Analytics in Healthcare: Key Benefits and Use Cases | Mercury Healthcare. [online] Available at: https://www.mercuryhealthcare.com/blog/predictive-analytics-healthcare.

5. Patel, N. (2022). 10 Predictive Analytics Use Cases &Examples In Healthcare Industry. [online] Make An App Like. Available at: https://makeanapplike.com/10-predictive-analytics-use-cases-examples-in-healthcare-industry/ [Accessed 14 Apr. 2023].

6. Philips (2020). Predictive analytics in healthcare: three real-world examples. [online] Philips. Available at: https://www.philips.com/a-w/about/news/archive/features/20200604-predictive-analytics-in-healthcare-three-real-world-examples.html.

7. SearchBusinessAnalytics. (n.d.). 7 Top Predictive Analytics Use Cases: Enterprise Examples. [online] Available at: https://www.techtarget.com/searchbusinessanalytics/feature/Top-5-predictive-analytics-use-cases-in-enterprises.

8. SearchBusinessAnalytics. (n.d.). Predictive analytics in healthcare: 12 valuable use cases. [online] Available at: https://www.techtarget.com/searchbusinessanalytics/tip/Predictive-analytics-in-healthcare-12-valuable-use-cases.

9. SearchEnterpriseAI. (n.d.). What is Predictive Modeling? [online] Available at: https://www.techtarget.com/searchenterpriseai/definition/predictive-modeling.

10. Team, W.E. (2022). Predictive Modeling. [online] WallStreetMojo. Available at: https://www.wallstreetmojo.com/predictive-modeling/.

11. TrendHunter.com. (n.d.). 24 Examples of Predictive Technology. [online] Available at: https://www.trendhunter.com/slideshow/predictive-tech [Accessed 14 Apr. 2023].

